# Exploring the role of the potential surface in the behaviour of early warning signals

**DOI:** 10.1101/2022.08.12.22278717

**Authors:** Andrew Nugent, Emma Southall, Louise Dyson

## Abstract

The theory of critical slowing down states that a system displays increasing relaxation times as it approaches a critical transition. These changes can be seen in statistics generated from timeseries data, which can be used as early warning signals of a transition. Such early warning signals would be of value for emerging infectious diseases or to understand when an endemic disease is close to elimination. However, in applications to a variety of epidemiological models there is frequent disagreement with the general theory of critical slowing down, with some indicators performing well on prevalence data but not when applied to incidence data. Furthermore, the alternative theory of critical speeding up predicts contradictory behaviour of early warning signals prior to some stochastic transitions. To investigate the possibility of observing critical speeding up in epidemiological models we characterise the behaviour of common early warning signals in terms of a system’s potential surface and noise around a quasi-steady state. We then describe a method to obtain these key features from timeseries data, taking as a case study a version of the SIS model, adapted to demonstrate either critical slowing down or critical speeding up. We show this method accurately reproduces the analytic potential surface and diffusion function, and that these results can be used to determine the behaviour of early warning signals and correctly identify signs of both critical slowing down and critical speeding up.

## 1. Introduction

Complex systems that exhibit critical transitions, characterised by a sudden shift in the state of the system, can be found in fields ranging from climate change [1, 2] to finance [3, 4] to epidemiology [5, 6]. By their nature, critical transitions have a major impact on a system’s behaviour. Substantial effort has been devoted to identifying both generic and model-specific methods for detecting impending critical transitions. One such method is that of early warning signals (EWSs) [5, 7, 8, 9]: summary statistics which monitor features of a timeseries that are known to change as the system approaches a ‘tipping point’.

EWSs arise from the theory of critical slowing down (CSD). The key concept of CSD is that as a system approaches a critical transition caused by a zero-eigenvalue bifurcation it becomes slower to recover from small perturbations away from its steady state [7, 8]. CSD can be explained by the size of the dominant eigenvalue of the system, which decreases towards zero as the bifurcation is approached, leading to longer return times. Alternatively CSD can be understood through the shape of the potential surface, which becomes flatter around the steady state, indicating a weakening of the dynamic forces pulling the system back to equilibrium. This effect becomes apparent in timeseries data through an increase in variance and autocorrelation as the system approaches the bifurcation, as well as in the behaviour of various other statistical indicators.

One important application of EWSs is to understand when a disease is approaching the point at which it can cause a major outbreak. In the wake of the COVID-19 pandemic there has been renewed attention in developing techniques such as these for identifying possible future pandemics before they become uncontrollable. Conversely, it is also necessary to robustly detect infections of potential concern, while avoiding undue false alarms. To develop EWSs with such characteristics requires a detailed understanding of their behaviour in various different types of models and situations. This paper aims to develop our understanding of the underlying reasons for the variety of EWS behaviours we observe in different models and data.

Although originally applied to saddle-node bifurcations [7], CSD has also been observed in other situations in which systems undergo critical transitions without crossing a bifurcation [10, 11, 12], and for systems undergoing a Hopf bifurcation [13] and transcritical bifurcations [5, 9]. It is important to note that CSD theory is not applicable to all critical transitions. In particular CSD would not be expected prior to a purely stochastic transition between a system’s existing steady states [14]. Dakos *et al*. provide a detailed description of the various causes of critical transitions and in which of these CSD can be observed [10]. We focus on situations in which a critical transition is induced by a slow change in some underlying parameter which itself approaches a ‘critical threshold’.

We return to the understanding of CSD as a flattening of the potential surface. Fig. 1 shows a section of a potential surface around a steady state, located at the local minimum of the potential surface. Four features of the potential surface are marked on the figure, corresponding to different measures of the system’s resilience [10]. *h* describes the height of the potential surface at its local minimum. *b* describes the distance to the border of the basin of attraction, that is the distance from the local minimum of the potential surface to the nearest local maximum. These two features approximate the distance to the system’s tipping point in parameter space and state space respectively. *ρ* describes the ratio between these (*ρ* = *h/b*) as an approximation to the steepness of the potential surface, while *λ* describes the steepness (second derivative) of the potential surface at its local minimum.

**Figure 1:**
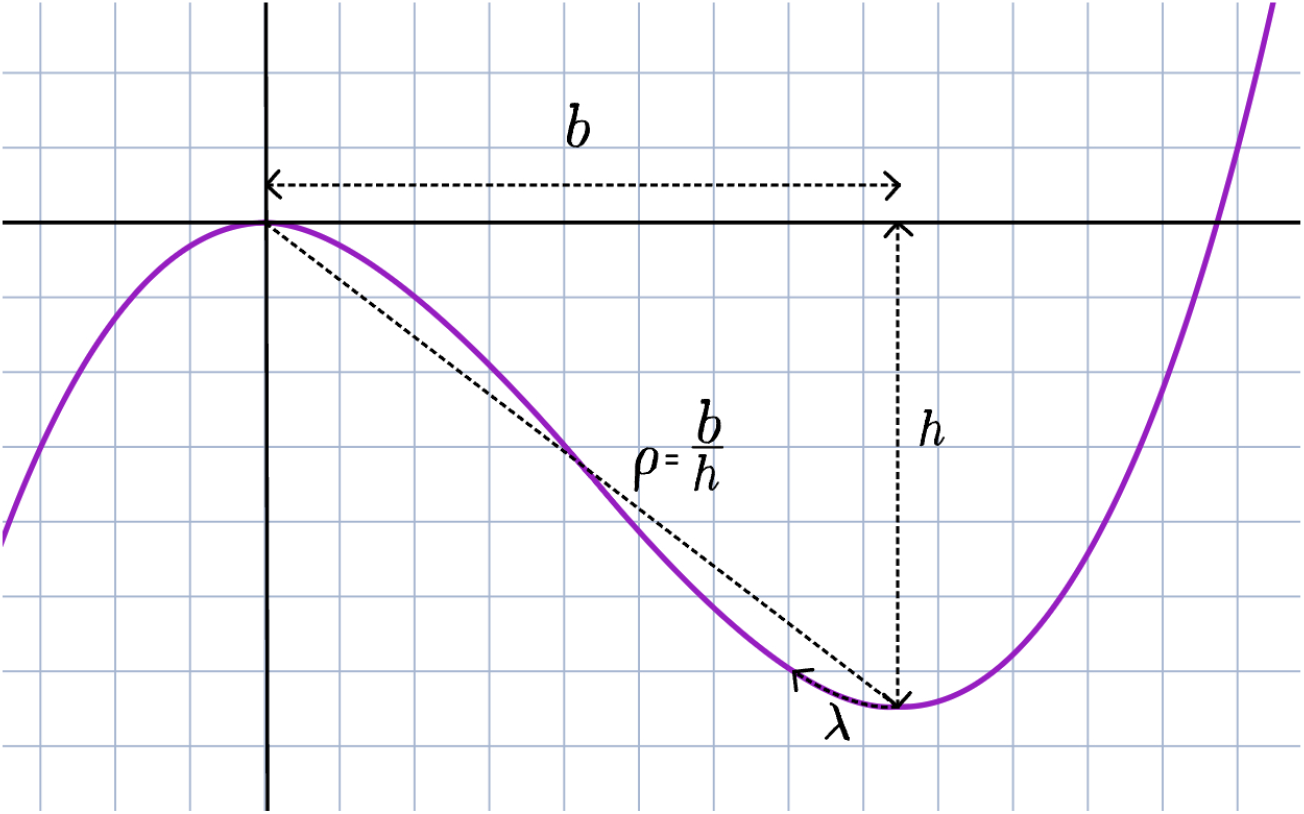
Four features of the potential surface are displayed: the depth of the surface at its local minimum (*h*), the distance to the border of the basin of attraction (*b*), the ratio of these (*ρ*), and the steepness of the potential surface at its local minimum (*λ*).

CSD is characterised by a shrinking of the basin of attraction and a flattening of the potential surface as the underlying parameter approaches its critical threshold. Hence *h, ρ* and *λ* would all be expected to decrease as the critical transition is approached.

CSD has been observed both in empirical data [15, 16] and in EWSs derived from analytic models [5, 17]. However there are also many cases in which the behaviour of EWSs does not match the statistical signatures associated with CSD in both theoretical models [17, 18, 19, 20, 21] and real data [3, 22, 23]. The expansion of EWSs to systems beyond standard bifurcation models, including stochastic systems in which a critical transition is made increasingly likely due to a slow parameter change [24], has added to the need for a more detailed understanding of the behaviour of EWSs.

Recently Titus and Watson proposed a possible explanation of unexpected EWSs behaviour through an alternative theory called critical speeding up (CSU) [24]. The key difference is that the potential surface is assumed to become steeper rather than flatter as the critical transition is approached. Referring to Fig. 1, this corresponds to an expected increase in *ρ* and *λ*, combined with a decrease in *b* and constant or decreasing *h*. As these changes occur in the shape of the potential surface, the system makes smaller, yet more frequent, excursions from the steady state until eventually leaving the narrowing basin of attraction. As with CSD the critical transition is approached by moving a model parameter towards a critical threshold, although in the case described by Titus and Watson the threshold cannot be attained as it requires a division by zero in the underlying model. The critical transition is not induced purely by noise, however, but also by the changing shape of the potential surface which results in such a transition becoming increasingly likely. The expected behaviour of EWSs are therefore reversed as the system becomes faster, not slower, to return to equilibrium. For example, variance and autocorrelation are both expected to decrease under CSU, whereas CSD predicts that they would both increase.

Although the concepts of CSD and CSU are clearly distinct and predict opposite EWSs behaviours, identifying which, if either, phenomena a system experiences may not be straightforward. Faced with EWSs whose behaviour deviates from the predictions of CSD there are various possible explanations [9, 10, 25]. Issues may arise from the calculation of EWSs [26], from the type of data used [25], from assumptions about the nature of the transition [11], or because a system experiences CSU rather than CSD [24]. In order for EWSs to provide meaningful information it is vital to have a good understanding of how these signals are expected to behave in the context of a particular application.

Here we present a situation-specific method for understanding the behaviour of EWSs. This approach, which we refer to as an ‘equation-free method’ (EFM) operates by reconstructing the potential surface and noise process from timeseries in which the underlying parameter is fixed [27, 28]. EWSs can be approximated from the features of the potential surface and noise at steady state. The chosen parameter value is then changed and the process is repeated to give an understanding of how EWSs change as the system moves towards the critical transition. This approach provides fresh insight into the complex behaviour of EWSs while also giving valuable information about the changing shape of the potential surface and its impact on a system’s dynamics. We demonstrate the EFM on a case study, which we adapt from a classic epidemiological model, designed to demonstrate either CSD or CSU depending on the choice of a fixed model parameter *n*. This case study shows how the behaviours of EWSs may deviate from the typical patterns associated with CSD and CSU, and how the EFM can be used to explain why these behaviours occur. We also discuss the applicability of the EFM to experimental data: while it offers a framework for calculating EWSs on unevenly spaced data and without requiring moving averages or data detrending, both of which pose problems when calculating EWSs directly from timeseries, it requires large volumes of data and is not proposed as a means to calculate EWSs in real time.

## 2. Methods

To clarify the relationship between the behaviour of the potential surface and the behaviour of EWSs, we derive analytic expressions for five common EWSs in terms of features of the potential surface and the system’s noise.

As in [24] we assume that the dynamics of a stochastic process *X* are controlled by a smooth potential function *V* plus some noise, modelled by the stochastic differential equation

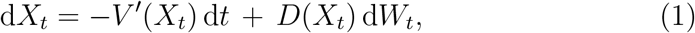

where (*W*_*t*_ : *t* ≥ 0) is a standard Weiner process. We assume that this system has a stationary or quasi-stationary state denoted 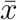, defined as a local minimum of *V* (the potential, with 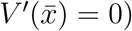), around which *X* fluctuates. By considering the linearisation of the dynamics around 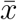 (for example by linearising in the corresponding Fokker-Planck equation), we obtain an Ornstein-Uhlenbeck process describing the fluctuations 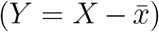 around this point [17, 18]. We denote the steepness of the potential surface at the steady state by 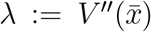 (as in Fig. 1) and the value of the diffusion function at the steady state by 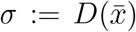, giving the following stochastic differential equation for the fluctuations

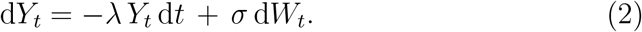

Analytic expressions for various statistics commonly used as EWSs are already established for this simpler process [29] and are displayed in Table 1. These expressions are given solely in terms of the steepness of the potential surface (*λ*), the value of the diffusion function at the fixed point (*σ*) and the location of the fixed point 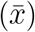. Note that they do not rely on other features of the potential surface, such as the precise width and depth of the basin of attraction. The expressions in Table 1 correspond to the behaviour of EWSs calculated on a system at its steady state. A similar approach to deriving the behaviour of EWSs is taken in [17] and [24].

**Table 1:** Common EWSs expressed in terms of the steepness of the potential surface at the fixed point (*λ*), the value of the diffusion function at the fixed point (*σ*), and the location of the fixed point 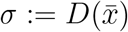.

| Early Warning Signal | General Form |
| --- | --- |
| Variance | $\frac{\sigma^2}{2\lambda}$ |
| Coefficient of variation | $\frac{\sigma}{\bar{x}\sqrt{2\lambda}}$ |
| Index of dispersion | $\frac{\sigma^2}{2\lambda\bar{x}}$ |
| Lag-1 autocorrelation | $e^{-\lambda}$ |
| Decay time | $\frac{1}{\lambda}$ |

**Table 2:**
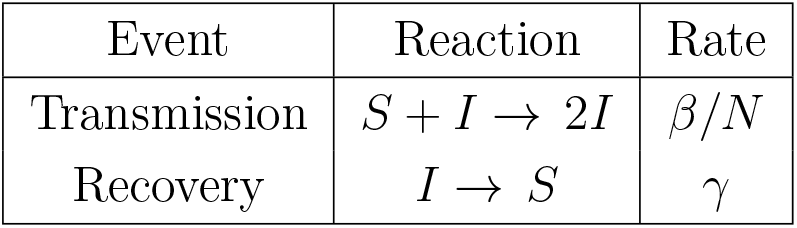
SIS model reactions.

As underlying parameters are varied the values of *λ, σ* and 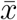 may also change. For a system that experiences CSD, characterised by a flattening of the potential surface, *λ* will decrease. This leads to the increase in autocorrelation that is typically associated with CSD. For systems experiencing CSU the opposite effect occurs: as *λ* increases, the autocorrelation decreases.

If *σ* is assumed to remain constant then CSD and CSU will lead to the expected increase and decrease in variance respectively. However, in models with multiplicative noise and in real-world complex systems, *σ* could increase or decrease as the critical transition is approached [17]. From this we see that the potential surface alone is insufficient to fully understand the behaviour of EWSs, as many also depend significantly on *σ*.

If the drift and diffusion functions (− *V* ^*′*^(*X*) and *D*(*X*) respectively) in Eqn. 1 are known then these can be used to compute 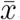, *λ* and *σ* directly. In the case that these equations are not known we present an ‘equation-free’ method to approximate the values of 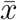, *λ* and *σ* from timeseries data and therefore understand the behaviour of various EWSs. The following method is adapted from [27, 28]. A similar method has also been presented in [30] and [31], the latter of which also describes an R package for their implementation.

We begin by taking an Euler-Maruyama discretisation [32] to approximate Eqn. 1 in discrete time-steps of length *δ*, giving

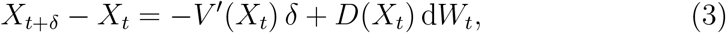

with d*W*_*t*_ = (*W*_*t*+*δ*_ − *W*_*t*_). For a specific value *x* in the range of the process, assume *X*_*t*_ = *x* and let *x*_*δ*_ be the position of the process at time *t* + *δ*. We then rearrange Eqn. 3 and average over many realisations (denoted by ⟨· ⟩) to obtain

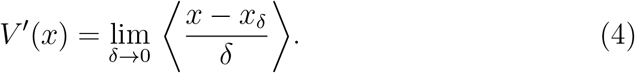

As d*W*_*t*_ has an expectation of zero, this part of the SDE vanishes in the average. By repeating this process across the range of *X* we obtain an approximation to the function *V* ^*′*^(*x*). We then integrate with respect to *x* to obtain the function *V*. This method is applied to timeseries data by partitioning the range of the data into sections and approximating *V* ^*′*^(*x*) in each section, before linearly interpolating and integrating numerically. These reconstructed potential surfaces can then be used to approximate 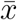 as well as features of the potential surface *b, h, ρ* and *λ*.

A visual representation of the implementation of this method is provided in the supplementary material Section 1.

We may approximate the diffusion function *D*(*X*) in a similar way. Squaring Eqn. 3 gives

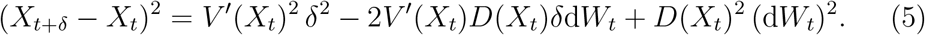

Averaging both sides of this equation over many realisations as before, then dividing by *δ* gives

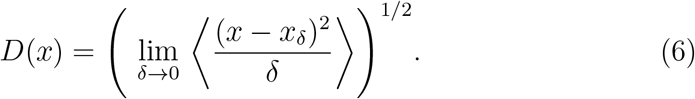

In practical computation, where taking *δ* → 0 is not possible and an approximation to *V* ^*′*^(*x*) has previously been calculated, an approximate form for a small fixed *δ >* 0 is given by

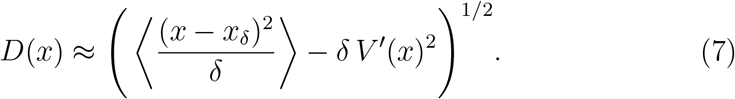

This provides an approximation to the value of the diffusion function at the fixed point 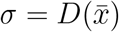. Using the forms in Table 1, the behaviour of the EWSs can now be obtained.

We demonstrate an application of this method to simulated timeseries for an adapted SIS model, capable of displaying either CSD or CSU. During each realisation of the simulation the parameter values are fixed, allowing us to reconstruct the potential surface and diffusion function for these set parameters. We can then investigate how the shape of the potential surface and behaviour of EWSs changes as the critical transition is approached by comparing the results between the different parameter values.

## 3. Case Study: Adapted SIS Model

The standard SIS model without demography can be described by the following reactions, where *S* represents one susceptible individual and *I* represents one infected individual.

The total population size (*N* = *S* + *I*) remains constant as no new individuals enter or leave the system, individuals only change their classification. These two reactions lead to the following SDE, derived in the supplementary material Section 2, where 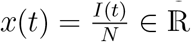 is the proportion of the population that is infected at time *t*,

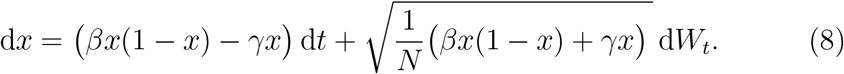

We refer to the terms preceding d*t* and d*W*_*t*_ as the drift *F* (*x*) and diffusion *D*(*x*) functions respectively,

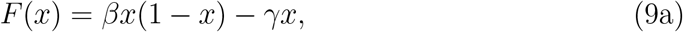

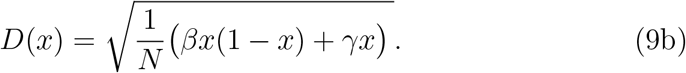

The mean field equation returns the standard non-dimensionalised ODE for the SIS model [33, 34],

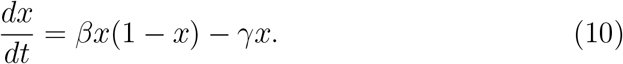

While the shape of the potential surface is determined by the parameters *β* and *γ*, the true parameter of interest is the basic reproduction number given by 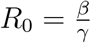. The system undergoes a transcritical bifurcation at *R*_0_ = 1 when the endemic steady state 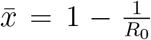 collides with the disease-free steady state, which becomes stable [35].

To construct a model that demonstrates a variety of behaviours corresponding to CSD and CSU we first consider *β* and *γ* as some specified functions of *R*_0_. To maintain the definition of *R*_0_ we require these functions to satisfy the relationship *β*(*R*_0_) = *R*_0_ *γ*(*R*_0_), so need only specify the form of *γ*(*R*_0_).

Titus and Watson propose that the difference between CSD and CSU is determined by the relationship between the width and depth of the basin of attraction [24]. As the width of the basin of attraction depends only on *R*_0_, and not on *β* and *γ* individually, we consider the depth of the potential surface,

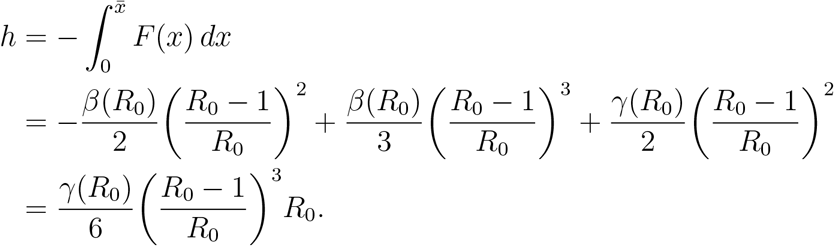

Based on this we choose *γ*(*R*_0_) of the form:

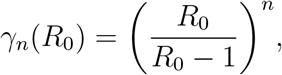

for values of *n* = 0, 1, 2, 3. Here *n* acts as a parameter that controls the behaviour of the system on the approach to the critical transition. For higher values of *n* the rate at which the depth of the basin of attraction decreases as *R*_0_ ↓ 1 is slower, indicating the possibility of observing CSU. Moreover, this form provides a potential surface with decreasing steepness for *n* ≤ 1 and increasing steepness for *n >* 1 (as *R*_0_ ↓ 1). Further features of this potential surface are given in Table 3.

**Table 3:** Features of the potential surface and noise process for the adapted SIS model. 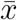: the position of the endemic quasi-steady state. *h* : the depth of the potential surface at 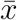. *b*: the distance from 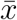 to the nearest edge of the basin of attraction. *ρ*: the ratio between *h* and *b*, approximating the steepness of the potential surface. *λ*: the steepness (second derivative) of the potential surface at 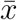. *σ*^2^: the value of the diffusion function at 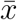 (squared). ↘ indicates a variable is decreasing as *R*_0_ decreases towards 1, → indicates it is constant and ↗ indicates it is increasing. Entries for which variables are constant or increasing are highlighted with a grey background.

| Feature | Expression | Trend as $R_0 \downarrow 1$ | | | |
| --- | --- | --- | --- | --- | --- |
| | | $n = 0$ | $n = 1$ | $n = 2$ | $n = 3$ |
| Endemic steady state ($\bar{x}$) | $1 - \frac{1}{R_0}$ | ↘ | ↘ | ↘ | ↘ |
| Potential surface depth ($h$) | $\frac{R_0}{6K_n} \left( \frac{R_0}{R_0 - 1} \right)^{n-3}$ | ↘ | ↘ | ↘ | ↘ |
| Distance to disease-free state ($b$) | $1 - \frac{1}{R_0}$ | ↘ | ↘ | ↘ | ↘ |
| Ratio ($\rho = h/b$) | $\frac{R_0}{6K_n} \left( \frac{R_0}{R_0 - 1} \right)^{n-2}$ | ↘ | ↘ | ↘ | ↗ |
| Potential surface steepness ($\lambda$) | $\frac{R_0}{K_n} \left( \frac{R_0}{R_0 - 1} \right)^{n-1}$ | ↘ | ↘ | ↗ | ↗ |
| Noise at endemic steady state ($\sigma^2$) | $\frac{2}{NK_n} \left( \frac{R_0}{R_0 - 1} \right)^{n-1}$ | ↘ | → | ↗ | ↗ |

To allow a direct comparison between different values of *n* we establish a starting value of *R*_0_ = 1.5 from which *R*_0_ is decreased towards 1, then normalise *γ*_*n*_ and *β*_*n*_ so that their values at *R*_0_ = 1.5 are independent of *n*. We define the normalising factor *K*_*n*_ = 3^*n*^ and

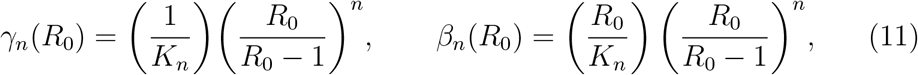

so that for all values of *n, γ*_*n*_(1.5) = 1, *β*_*n*_(1.5) = 1.5.

In this model the exhibited behaviour (CSD/CSU) will be controlled by the value of *n*. We note that a result of requiring that *β*_*n*_(*R*_0_)*/γ*_*n*_(*R*_0_) = *R*_0_ is that if time is rescaled by the recovery rate, then the model dependence on *n* is removed and the dynamics correspond to the SIS model. While this model provides a useful tool to study CSD and CSU, their underlying causes and the differences between them, we do not assert that this represents any particular real-world system, nor that a system exists in which changing a given parameter would move the behaviour from CSD to CSU.

The stochastic differential equation for disease prevalence (*x* ∈ ℝ) is now

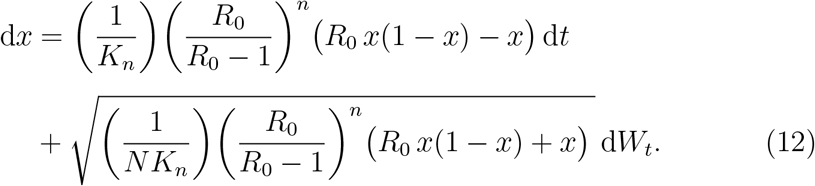

For *n* = 0 this model corresponds to the standard SIS model in Eqn. 8. For *n >* 0 the system no longer bifurcates, instead as *R*_0_ is slowly reduced towards 1 the system is driven towards a stochastic transition to the disease free state. Despite this the system exhibits CSD for both *n* = 0 and *n* = 1. For *n* = 2 and *n* = 3 the model instead exhibits signs of CSU. We do not consider values *n* ≥ 4, as this leads to the depth of the potential surface growing to infinity as *R*_0_ approaches 1. For all values of *n* considered the critical transition is induced by the reduction of the parameter *R*_0_ towards its critical threshold.

The potential surface for the adapted SIS model is found by integrating the negative of the drift function Eqn. 9a and substituting the functions *β*(*R*_0_) and *γ*(*R*_0_) to give

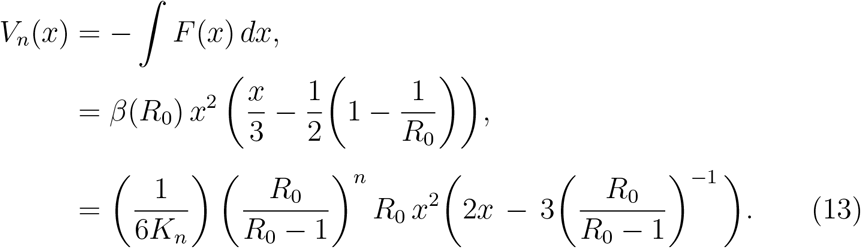

From Eqn. 12 and Eqn. 13 we analytically calculate features of the potential surface and noise at the endemic steady state. These results are summarised in Table 3.

The steepness of the potential surface (*λ*) is increasing for *n* = 0, 1 and decreasing for *n* = 2, 3. This indicates that the system experiences CSD for *n* = 0, 1 and CSU for *n* = 2, 3. We refer to this change as the CSD/CSU threshold for *n*. The behaviour of *σ* is notably different from *λ*, decreasing for *n* = 0 only. This highlights the fact that trends in the noise of the process may not match trends in the steepness of the potential surface as the critical transition is approached, hence it is vital to understand these two features separately.

The location of the steady state, 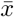, is decreasing for all values of *n*. This indicates that the basin of attraction is becoming narrower, but does not provide an indication of CSD or CSU. Similarly, a decrease in *h* and *b* is observed for all values of *n*, again indicating that the system is approaching a critical transition, but this trend also provides no indication of whether the critical transition will be preceded by CSD or CSU. *ρ* is decreasing for *n* = 0, 1, 2 and increasing for *n* = 3, so its change in behaviour does not correspond to the CSD/CSU threshold. Hence an increasing or decreasing trend in *ρ* is not a clear indicator of CSU.

The trends seen in the features of this model further motivate the focus on recovering values of 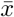, *λ* and *σ* as a means to understand the behaviour of a system on the approach to a critical transition.

Substituting the analytic expressions above for 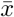, *λ* and *σ* into the general forms given in Table 1 we obtain analytic expressions for the behaviour of EWSs for this adapted SIS model, presented in Table 4. These expressions match those obtained using the linear noise approximation performed on the SIS model by Southall *et al*. [9] when the system is at the endemic fixed point, substituting *β*(*R*_0_) and *γ*(*R*_0_) where needed.

**Table 4:** Analytic expressions of five common EWSs applied to the adapted SIS model for general *n*. These expressions are derived by substituting the values of *λ, σ* and 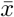 obtained from the analytic SDE into the general form of the EWSs given in Table 1.

| Early Warning Signal | Analytic Expression |
| --- | --- |
| Variance | $\frac{1}{R_0}$ |
| Coefficient of variation | $\frac{\sqrt{R_0}}{R_0 - 1}$ |
| Index of dispersion | $\frac{1}{R_0 - 1}$ |
| Lag-1 autocorrelation | $\exp\left(-\frac{R_0}{K_n}\left(\frac{R_0}{R_0 - 1}\right)^{n-1}\right)$ |
| Decay time | $\frac{K_n}{R_0}\left(\frac{R_0}{R_0 - 1}\right)^{1-n}$ |

We now present the results of the EFM applied to the adapted SIS model. For each value of *n*, the potential surface and diffusion function were reconstructed for five values of *R*_0_, decreasing towards *R*_0_ = 1. The values of 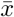, *λ* and *σ* were then approximated to give insight into the overall behaviour of the system (CSD/CSU/neither) as well as the behaviour of EWS (increasing/decreasing/neither).

For each value of *n* the five potential surfaces are plotted in Fig. 2 to demonstrate how the shape of the surface changes as *R*_0_ decreases from 1.5 to 1.1. For all *n* values the potential surfaces plotted for *R*_0_ = 1.5 (front-most potential surface in each figure) are identical. Fig. 2 shows how the potential surface becomes visibly flatter as *R*_0_ decreases for *n* = 0, 1 and visibly steeper for *n* = 2, 3. Plots of the analytic equation for the potential surface are given in red dashed lines for comparison. Solid blue lines show the results of the EFM applied for each combination of *n* and *R*_0_. In all cases there is an excellent fit between the analytic equation and the potential surface from the EFM.

**Figure 2:**
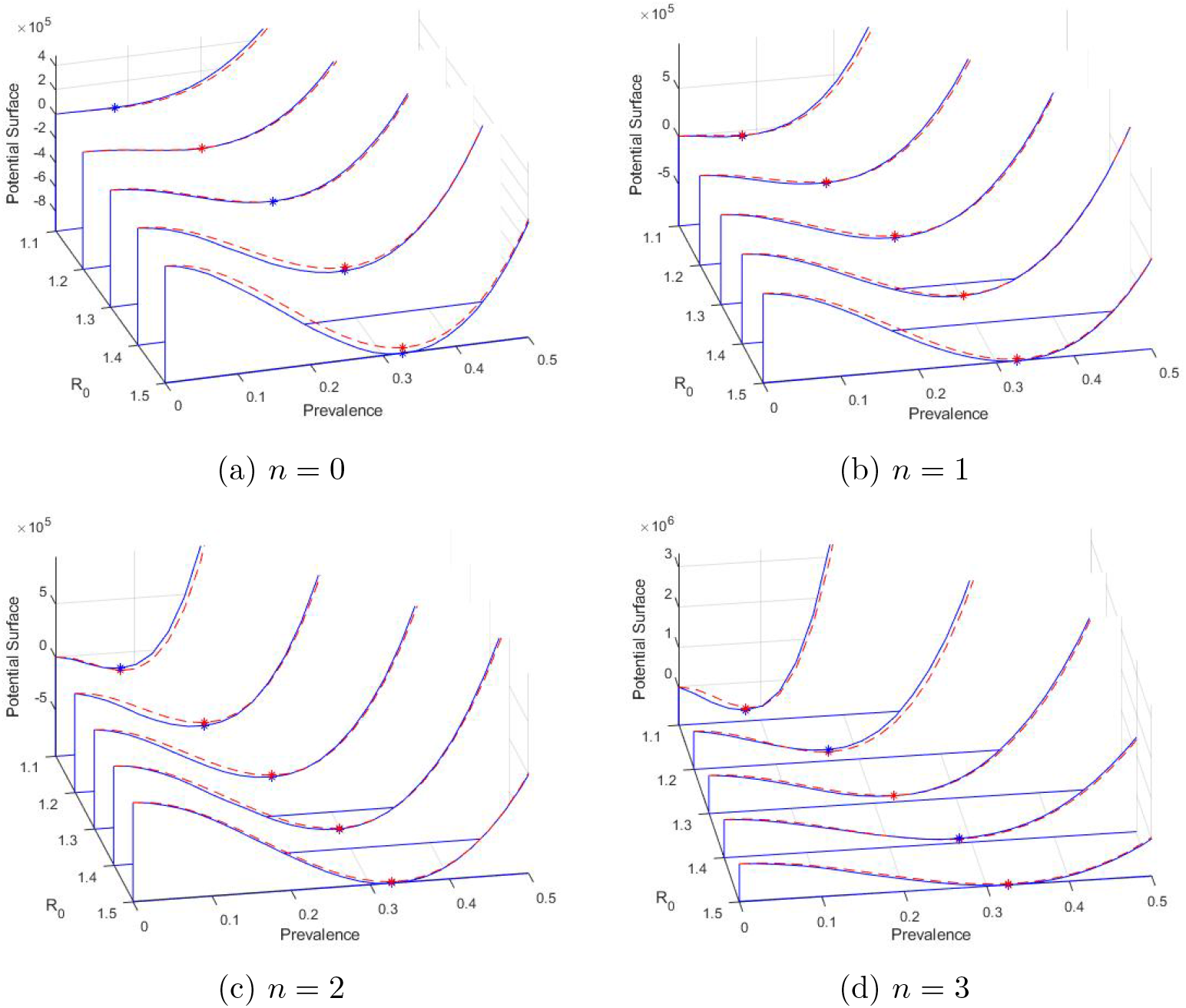
Comparing analytic (red dashed) and EFM (blue) results for the potential surface for the adapted SIS model. There is an excellent fit between the two sets of results, hence the EFM could be used to take approximate measures of the depth, width and steepness of the potential surfaces. 100 simulations have been run for each *R*_0_ and *n* value. (a) Comparing analytic expressions for **autocorrelation** against results from the EFM (on the left) and simulations in which *R*_0_ is slowly varied (on the right). Both sets of numerical results match the analytic expressions well. (b) Comparing analytic expressions for **variance** against results from the EFM (on the left) and simulations in which *R*_0_ is slowly varied (on the right). Both sets of numerical results match the analytic expressions well.

The steady state, 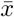, can be found by locating the minimum of the potential surface. *λ* was then estimated from the EFM by fitting a polynomial function to a portion of the potential surface and calculating the steepness of the polynomial at 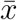. The EFM was also successful in accurately approximating the drift function *D*(*x*). This approximation was then used to estimate 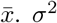 for each pairing of *R*_0_ and *n* values.

We now have three methods for calculating EWSs for this model: the first method is the analytic expressions given in Table 4; the second method is to substitute the approximations of 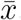, *λ* and *σ* from the EFM into the general forms in Table 1; the third method is to calculate EWSs directly on simulated timeseries in which *R*_0_ is slowly decreased. In these timeseries *R*_0_ is a linear function of time, given by

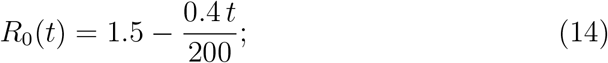

for *t* ∈ [0, 200], and to calculate EWSs we take outputs at integer intervals. A comparison of the results of these three methods is given in Fig. 3.

**Figure 3:**
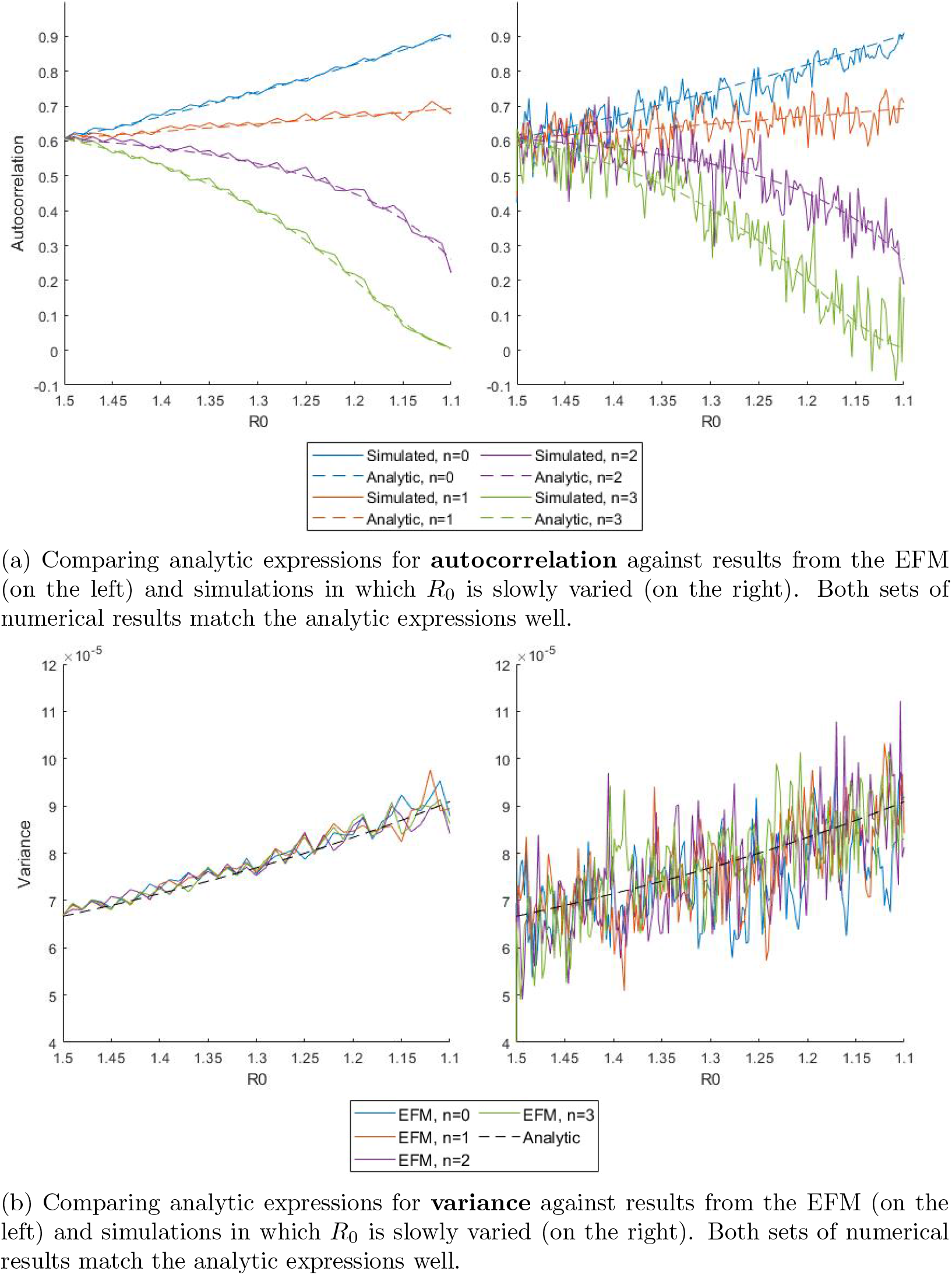
The behaviour of lag-1 autocorrelation (a) and variance (b) for the adapted SIS system. Note that the figures on the right show results at significantly more values of *R*_0_.

In the left column of Fig. 3 we compare analytic results (method one) with results from the EFM (method two). In the top-left we show the behaviour of lag-1 autocorrelation (Fig. 3a) and in the bottom-left the behaviour of variance (Fig. 3b), with a clear match between the two methods in both cases. This demonstrates the success of the EFM in approximating 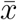, *λ* and *σ* and accurately predicating the behaviours of these EWSs.

In the right column of Fig. 3 we compare analytic results (method one) to the calculation of EWSs on simulated timeseries in which *R*_0_ is slowly decreased (method three). We refer to these as ‘time-varying parameter’ timeseries, whereas the EFM uses ‘fixed parameter’ timeseries. For each of 200 Gillespie simulation realisations the value of *R*_0_ is slowly decreased from 1.5 to 1.1. EWSs are calculated between the realisations at each timepoint. Again there is a good match between the two methods. It should be noted that the time-varying parameter simulations pass through many more values of *R*_0_ than the EFM, leading to the comparison with analytic results appearing noisier. Overall our results indicate that both the analytic expressions and the EFM results accurately represent the behaviour of EWSs as the system approaches a critical transition. Corresponding results for all EWSs are given in the supplementary material Section 3.

## 4. Discussion

There are two main conclusions that can be drawn from this case study: firstly, the EFM was successful in approximating the potential surface, diffusion function and behaviours of EWSs; and secondly, the behaviours of EWSs may deviate significantly from the typical patterns associated with CSD and CSU.

A striking feature of the expressions for the five common EWSs presented in Table 4 is that three (variance, coefficient of variation and index of dispersion) are *n*-independent. This behaviour is supported by the results in Fig. 3 of the EFM and simulations with time-varying *R*_0_. According to CSD/CSU theory all of variance, coefficient of variation and index of dispersion should increase for *n* = 0, 1 and decrease for *n* = 2, 3. This apparent contradiction can be explained by investigating the expressions for the EWSs given in Table 1, where their behaviour is explained in terms of 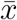, *λ* and *σ*. If the diffusion function is assumed to be constant then the change in *λ* leads to the expected increase (CSD) and decrease (CSU) in variance [7, 24]. However, we present here a model where the change in the shape of the potential surface is counterbalanced exactly by the change in the level of noise around the fixed point, leading to *n*-independence in these three EWSs. We conclude that, without additional information describing the noise around the fixed point, the behaviour of variance as an EWSs cannot be determined by the presence of either CSD or CSU, and cannot be used to distinguish between these phenomena. A similar conclusion can be reached for the coefficient of variation and index of dispersion. This highlights the importance of including realistic multiplicative noise when modelling any system with the goal of studying EWSs. This also demonstrates the usefulness of the EFM: by approximating 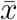, *λ* and *σ* the behaviour of EWSs are explained as well as calculated.

As the behaviour of autocorrelation, and therefore decay time, depends only on *λ* these can be determined by the presence of CSD or CSU alone, and can be considered characteristic of these phenomena. One limitation of autocorrelation as an EWS is the need for evenly spaced data points to be able to calculate autocorrelation either over a moving window or between realisations. Interpolating existing timeseries in which data points are not evenly spaced can introduce artificial autocorrelation [25], reducing the reliability of autocorrelation as an EWS. The EFM offers an alternative method for calculating autocorrelation from timeseries data, as there is no requirement for evenly spaced data. Eqn. 4 involves the average direction of travel of a timeseries from a given point, but does not require that the timeseries data be evenly spaced. Results shown here for the EFM can also be obtained using timeseries with unevenly spaced data.

The EFM may also offer a solution to other problems arising when calculating EWSs directly on timeseries data. As discussed by Boettiger and Hasting [14] a system may have multiple stable states and a timeseries may move between these. Calculating EWSs on this timeseries over a moving window could show increasing variance and autocorrelation preceding the movement between steady states. This would not be an occurrence of CSD as there has been no parameter change prompting this transition, the system is simply moving around its existing steady states due to the effects of stochasticity. Observing only the EWSs calculated on the timeseries, such a situation may be misinterpreted as it appears to confirm the theory of CSD. As the EFM reconstructs the drift function at each point in the specified range, multiple fixed points within this range could be identified. The behaviour of EWSs at a particular fixed point is then established by changing the parameter of interest and repeating the EFM, for example in Fig. 2 the EFM is repeated for five different *R*_0_ values decreasing from 1.5 to 1.1. The EFM therefore ensures that changes in EWSs are caused solely by changes to the underlying parameters, rather than other influences within the timeseries.

The calculation of EWSs directly on timeseries is also made more challenging by the need to detrend data, since the method chosen can then impact the reliability of the EWSs [26]. When only a single timeseries is used it is also necessary to calculate EWSs over a moving window. This relies on the additional assumption that a timeseries is ergodic, and thus that calculating EWSs over a moving window is equivalent to calculation over multiple realisations. This assumption, as well as the necessity to choose a moving window length, raises additional difficulties [25]. While the EFM offers an alternative approach it does not resolve these issues. The EFM method requires a large volume of data and is not suitable for calculating EWSs on a single timeseries. The challenges of detrending and ergodicity are reduced when multiple realisations are available, and in these circumstances, the EFM can be used to approximate the potential landscape; offering additional benefits for understanding the overall system and thus understanding the behaviour of EWSs.

It should be noted that the EFM is not proposed as a method for calculating EWSs and detecting critical transitions in real time. This is partly due to the requirement for fixed parameter timeseries, which would not be available when monitoring a real system believed to be approaching a critical transition. Additionally, multiple timeseries with varying initial conditions are typically required for each parameter value to allow the system to fully explore its potential surface. Furthermore, there must be sufficient data for the averages within the EFM to accurately represent the drift and diffusion functions. Therefore the EFM is best suited to systems which can be simulated, or where experimental data is available for given parameter values. To satisfy the assumptions of the EFM the underlying system must be a stationary Markov process for each parameter value, as is the case in the example provided here.

One drawback of the EFM is the need to select parameter values for which the potential surface and diffusion function should be constructed. In this case study developed from the SIS model and in other epidemiological situations, a critical transition commonly occurs at *R*_0_ = 1, with the precise definition of *R*_0_ varying between models. In broader applications identifying the parameter of interest and the value at which a critical transition may occur requires some prior understanding of the particular system.

There are various means by which investigation of the EFM could be expanded. The case study presented here is a one-dimensional system. Many complex systems, such as highly interconnected ecosystems, require models in multiple dimensions. While the dynamics of some can be reduced, expanding the EFM into multiple dimensions may improve prospects for its applicability. It is worth noting that since the domain of each variable must be divided into sections in which the drift and diffusion functions are approximated, the computational cost of the EFM would grow exponentially in the number of dimensions, likely making it impractical for high-dimensional systems.

The methodology of the EMF may also be further developed. The EFM approximation of the drift function involves calculating the average direction of the travel of the system given its current position. In Eqn. 4 this is done by averaging the forward difference. Replacing this forward difference with a central difference we obtain Eqn. 15,

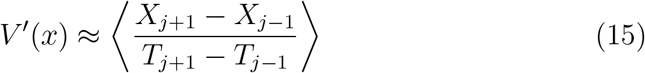

where *X* is the vector of system states at times *T*, with *X*_*j*_ = *x*. This adaptation to the EFM also accurately approximated the behaviour of the potential surface, diffusion function and EWSs for evenly and unevenly spaced data, but the full impact of such alterations is not known.

The EFM is proposed primarily as a route to understanding how a system changes as a critical transition is approached and therefore how EWSs are expected to behave. There are various examples of systems in which the behaviour of EWSs, especially variance, calculated on time-varying parameter data deviates from the typical pattern of CSD. It is crucial to understand why this occurs to ensure EWSs can be interpreted correctly. Is there some issue arising from the method of calculation? Does the system perhaps experience CSU rather than CSD? Does the nature of the system’s stochasticity also change as the critical transition is approached? The EFM offers an approach through which these highly interconnected questions can be separated. When monitoring real time data for signs of an impending critical transition practical calculation and interpretation issues still remain, yet the EFM provides a route to a clearer understanding of how the dynamics of a particular system change and how EWSs should be interpreted.

## Data Availability

Model code is available online at https://github.com/AJNugent1/CSD-CSU-Paper

## Code availability

MATLAB code for this paper may be found at https://github.com/AJNugent1/CSD-CSU-Paper.

