## Supplementary Information for "Exploring the role of the potential surface in the behaviour of early warning signals"

#### 1 Implementation of the EFM

We describe here the general method for implementing the EFM on timeseries data for a fixed parameter value. Full code can be found at `https://github.com/AJNugent1/CSD-CSU-Paper`.

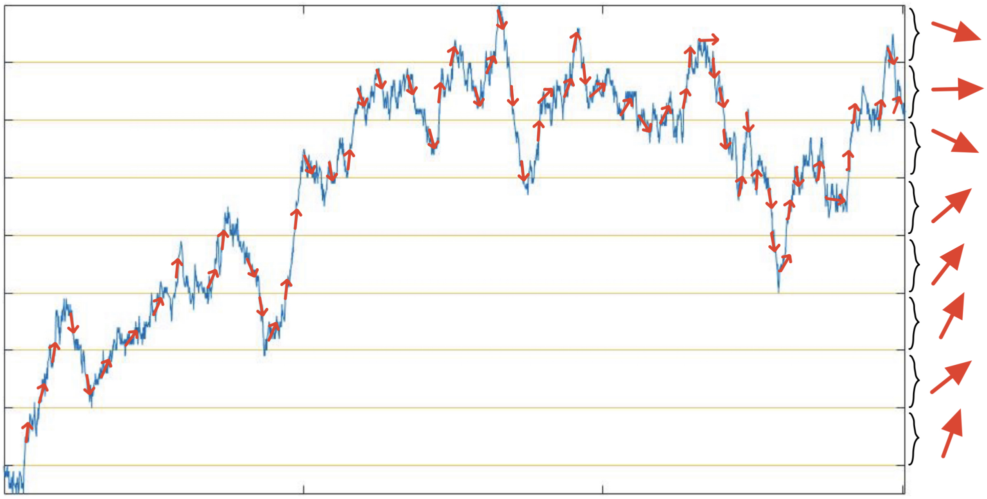

Figure 1: Visualisation of the EFM. An example timeseries is shown in blue. Yellow lines show the partition of the range of the timeseries into sections. Red arrows on the timeseries shown approximations to the drift function at various points, while larger red arrows on the right of the figure show the average of these approximations within each section. It is these averages that give the approximation of the drift function from the EFM.

1. Partition the range of the timeseries into intervals (shown in yellow in Fig. 1). The range covered by the intervals will determine where the drift and diffusion functions are constructed, this does not need to be the entire range of the timeseries.

2. We denote the timeseries by the pair $(T, X)$ of times $T$ and positions $X$, and individual points by $(T_k, X_k)$. For each $k$, identify which interval the position

$X_k$ lies in, then approximate the drift function and size of the diffusion function at this point using equations 4 and 6 or 7 from the main paper. In Section 2 in the main paper these approximations are calculated using $(X_{k+1}-X_k)/(T_{k+1}-T_k)$ and $(X_{k+1}-X_k)^2/(T_{k+1}-T_k)$ respectively, where $(T_{k+1}, X_{k+1})$ is the next point in the timeseries. Approximations to the direction of the drift function are shown as red arrows on the timeseries in Fig. 1. Note that the inclusion of $T_{k+1}-T_k$ accounts for unevenly spaced data points.

3. Average the approximated quantities over points within each section. This is shown in Fig. 1 by the larger red arrows on the right hand side. For the diffusion function we then take the square root. This gives an approximation to the values of the drift and diffusion function in each section, which can be taken either as a piecewise constant approximation or linearly interpolated. The potential surface is then given by the numerical integration of the drift function multiplied by -1.

Other comments on implementation of the EFM:

- Multiple timeseries are typically required to give enough data points in each section for the averages to accurately represent the drift and diffusion functions. Shorter timeseries beginning at different initial conditions are more informative than longer timeseries that remain nearby a single fixed point.

- Increasing the number of sections (by decreasing the width) provides greater detail for the drift and diffusion functions, at the cost of reduced accuracy as there are fewer data points within each section. The width of sections does not need to be equal, and can be tailored to focus on areas of interest or areas where more data is available.

- As mentioned in the Discussion, alternative approximations could be used for the drift function in step 2, such as the central or backwards difference.

### 2 Derivation of SDE for SIS model

In the following section we assume $R_0$ is fixed and denote by $\beta$ and $\gamma$ the values of the corresponding functions evaluated at our specified value of $R_0$. Recall the two model reactions and their rates:

| Event | Reaction | Rate |
|---|---|---|
| Transmission | $S+I \to 2I$ | $\beta/N$ |
| Recovery | $I \to S$ | $\gamma$ |

Table 1: SIS model reactions.

In order to derive a stochastic differential equation for the proportion of infected individuals, we follow the method taken in [1] for coarse graining the individual-based model above. Let $x$ denote the proportion of infected individuals at time $t$. Letting $T(a|b)$ be the rate of transition from state $b$ to state $a$, we have that the rate at which $x$ increases is given by

$T_+ := T\left(x + \frac{1}{N} \,|\, x\right) = \dfrac{\beta S I}{N} = \beta N x(1-x)$

and the rate at which $x$ decreases is given by

$T_- := T\left(x - \frac{1}{N} \,|\, x\right) = \gamma I = \gamma N x$

Define $P(x,t)$ as the probability density function of $x$ at time $t$. The transition rates above then give the master equation for $P(x,t)$ [2]:

$$\frac{\partial P}{\partial t}(x,t) = P\Big(x - \frac{1}{N}, t\Big)\,(T_+) - P(x,t)\,(T_+) + P\Big(x + \frac{1}{N}, t\Big)\,(T_-) - P(x,t)\,(T_-) \tag{1}$$

This PDE involves simultaneously $P(x,t)\,, P(x-\frac{1}{N},t)$ and $P(x+\frac{1}{N},t)$. To resolve this, we introduce the step operators $\epsilon^{\pm}$, which represent the creation or recovery of one infectious individual. We can represent $P(x \pm \frac{1}{N}, t)$ as $\epsilon^{\pm}P(x,t)$. For most elementary functions $f$, we can Taylor expand in $\frac{1}{N}$ [2]:

$$\epsilon^{\pm} f(x) = f\left(x \pm \frac{1}{N}\right) \approx \left(1 \pm \frac{1}{N}\partial_x + \frac{1}{2N^2}\partial_x^2\right) f(x) \tag{2}$$

Applying these step functions to our master equation, neglecting terms of order $N^{-3}$ or smaller, gives:

$$\begin{aligned}\frac{\partial P}{\partial t} &= (\epsilon^- - 1)P(x,t)\,T_+ + (\epsilon^+ - 1)P(x,t)\,T_- \\ &= \frac{1}{N}\frac{\partial}{\partial x}P(x,t)\,(T_- - T_+) + \frac{1}{2N^2}\frac{\partial^2}{\partial x^2}P(x,t)\,(T_- + T_+)\end{aligned}$$

Substituting $T_+$ and $T_-$, and re-scaling time $t \to \frac{1}{N}t$ gives our final result:

$$\frac{\partial P}{\partial t} = \frac{\partial}{\partial x}\Big(P(x,t)\,(\gamma x - \beta x(1-x))\Big) + \frac{1}{2N}\frac{\partial^2}{\partial x^2}\Big(P(x,t)\,(\gamma x + \beta x(1-x))\Big) \tag{3}$$

which is the Fokker-Planck equation for the corresponding SDE

$$dx = -(\gamma x - \beta x(1-x))\,dt + \sqrt{\frac{1}{N}\,(\gamma x + \beta x(1-x))}\;dW \tag{4}$$

### 3 Comparison of EWSs behaviours

For each of the EWSs in Table 4 of the main paper we compare the analytic expressions, results of the EFM and results of direct calculation on simulations in which $R_0$ is slowly decreased.

For figures on the left, potential surfaces and drift functions constructed using the EFM are used to give approximations to $\bar{x}, \lambda$ and $\sigma$ which are substituted into the general forms in Table 1 of the main paper to approximate the behaviour of EWSs. For the figures on the right, 500 Gillespie simulations are run in which the value of $R_0$ is slowly decreased towards 1 for each value of $n$. EWSs are calculated between the realisations. For both we compare against the analytic expressions in Table 4 of the main paper.

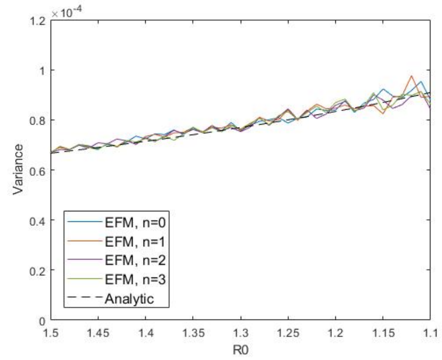


(a) Variance, EFM

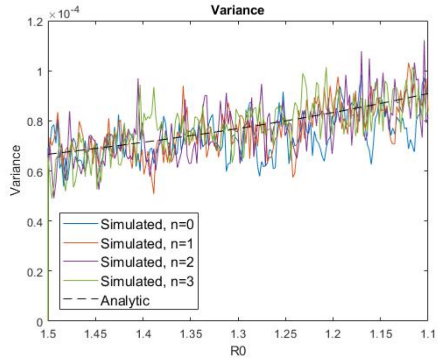


(b) Variance, simulated

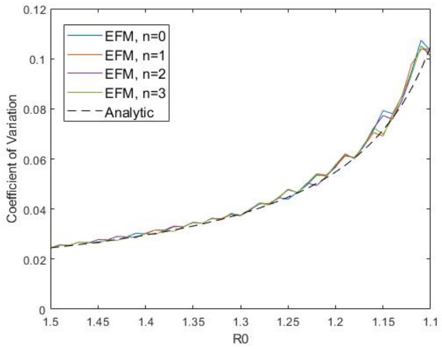


(c) Coef. of variation, EFM

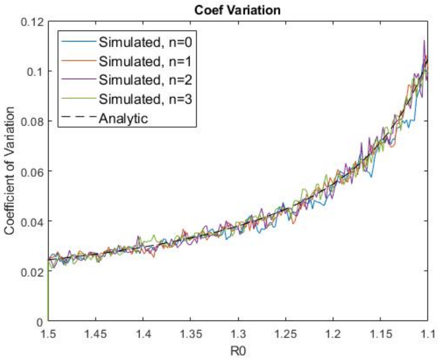


(d) Coef. of variation, simulated

Figure 2: Comparing analytic expressions for EWS against results from the EFM (on the left) and simulations in which $R_0$ is slowly varied (on the right). Both sets of numerical results match the analytic expressions well.

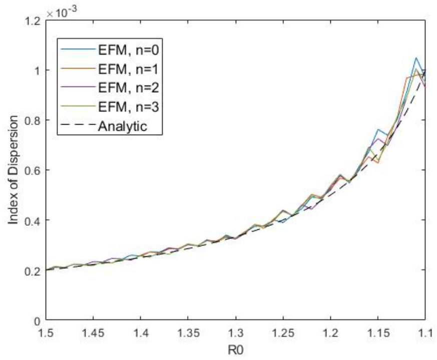


(a) Index of dispersion, EFM

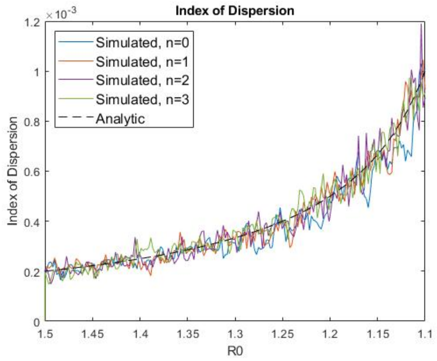


(b) Index of dispersion, simulated

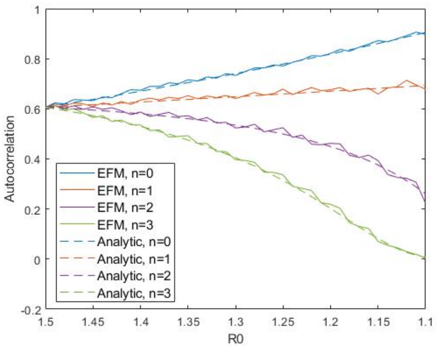


(c) Lag-1 autocorrelation, EFM

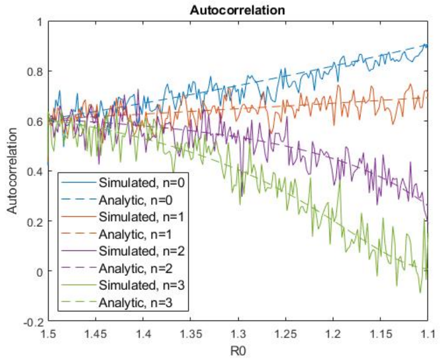


(d) Lag-1 autocorrelation, simulated

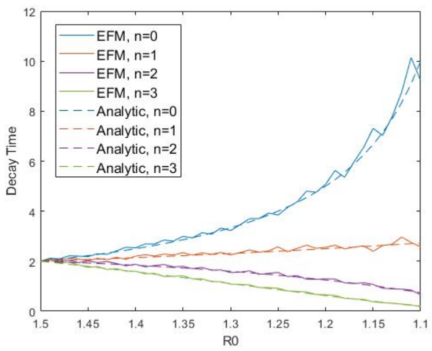


(e) Decay time, EFM

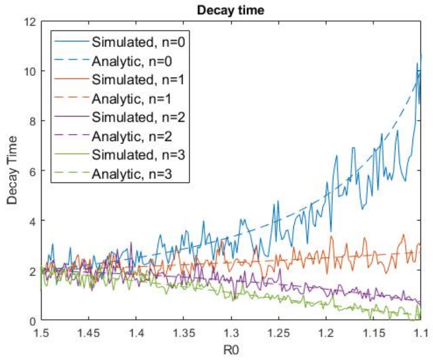


(f) Decay time, simulated

Figure 3: Comparing analytic expressions for EWS against results from the EFM (on the left) and simulations in which $R_0$ is slowly varied (on the right). Both sets of numerical results match the analytic expressions well.
